# The Role of Ethics in Physiotherapy: A Scoping Review Protocol

**DOI:** 10.1101/2024.10.28.24316250

**Authors:** Gianluca Bertoni, Sara Patuzzo Manzati, Federica Pagani, Marco Testa, Simone Battista

## Abstract

**Background and aims:** Ethical considerations play a crucial role in physiotherapy, influencing patient care, professional conduct, and clinical decision-making. Despite its significance, there is a limited comprehensive understanding of how ethical principles are applied in physiotherapy practice. The evolving nature of the field, alongside advancements in treatment approaches, presents new ethical challenges that require systematic investigation. This scoping review aims to map the existing literature on ethical issues within physiotherapy, identify research methodologies, and highlight knowledge gaps.

**Method:** This review will follow the methodological framework proposed by the Joanna Briggs Institute (JBI) for scoping reviews and will be reported following the PRISMA for Scoping Reviews guidelines. A comprehensive search will be conducted on PubMed, Medline, Embase, CINAHL, PsychInfo, Cochrane Central, and Pedro. The gray literature will be consulted. Studies involving physiotherapists and those addressing ethical issues in physiotherapy practice will be included. Data extraction will be based on a standardized form, and a narrative synthesis will categorize the ethical issues and principles.

**Discussion:** The review will provide a broad overview of ethical issues and principles in physiotherapy. It will inform future research priorities, guide ethical training for practitioners, and support the development of policies and guidelines to improve ethical shared decision-making in physiotherapy practice.

## 1. Introduction

In physiotherapy, ethical considerations are necessary to deliver high-quality care and ensure shared decision-making and positive outcomes for patients^1^. Physiotherapists frequently encounter ethical dilemmas such as balancing patient autonomy with professional recommendations^2^, managing conflicts of interest^3^, addressing disparities in access to care^2,4^, ensuring informed consent^5^, and navigating situations where the patient’s best interest may conflict with institutional policies or resources^6^. These dilemmas can significantly impact their practice and the patient experience^7–9^.

In light of the frequent necessity to navigate these ethical dilemmas, ethics plays a crucial role in physiotherapy^7^. However, the intersection of physiotherapy and ethics seems to have not received sufficient exploration, and our understanding of typical ethical issues in rehabilitation contexts is limited. We know little about the ethical principles adopted in physiotherapy and the philosophical disciplines or theoretical frameworks utilized in the literature to address these topics ^2,3,6,8–10^. Moreover, the evolving nature of physiotherapy practice, with advancements in treatment methods and a focus on patient-centered care, brings new ethical challenges that require careful consideration^7^. Hence, there is a need for a thorough examination of how these ethical issues are addressed in the literature.^7^. Moreover, the evolving nature of physiotherapy practice, with advancements in treatment methods and a focus on patient-centered care, brings new ethical challenges that require careful consideration^1,7,11^.

### 1.1 Rationale for conducting a scoping review

While the importance of ethical considerations in physiotherapy has been established, specific insights into applying these principles in practice still need to be explored. There is a pressing need to explore the unique ethical dilemmas physiotherapists face in various rehabilitation contexts and the principles and frameworks that guide their decision-making. A comprehensive understanding of how ethical challenges are addressed in different settings could inform the development of practical guidelines and training programs on the importance of applying ethics in clinical-decision making. By examining existing literature through a scoping review, we can identify key themes, gaps, and emerging issues related to ethics in physiotherapy. This exploration will enhance our understanding of ethical practices and provide a foundation for improving clinical practice, ultimately benefiting patient outcomes.

A scoping review is particularly suited for this investigation due to several reasons:

1. Broad Overview of Existing Knowledge: A scoping review will allow us to map the existing literature on ethics in physiotherapy, providing a broad overview of the key ethical issues, themes, and challenges that have been identified.
2. Identification of Gaps and Emerging Issues: By systematically examining the literature, a scoping review can highlight areas where research is lacking or where ethical challenges are not yet fully addressed. This will inform future research priorities and guide the development of targeted studies that address these gaps.
3. Integration of Diverse Perspectives: Physiotherapy ethics intersect with various aspects of clinical practice, including patient care, professional behavior, and institutional policies. A scoping review will integrate perspectives from different sources, including empirical studies, theoretical discussions, and practical guidelines, providing a comprehensive view of how ethics are integrated into physiotherapy practice.
4. Foundation for Policy and Practice Improvements: Understanding the current state of knowledge on ethics in physiotherapy will contribute to the development of better guidelines, training programs, and policies. It will support physiotherapists in navigating ethical dilemmas more effectively and ensure that ethical considerations are embedded in everyday practice.
5. Enhancement of Professional Development: By clarifying the ethical issues faced by physiotherapists and how they are managed, this scoping review will contribute to the professional development of practitioners. It will help enhance ethical awareness and decision-making skills among physiotherapists, ultimately improving the quality of patient care.

In summary, conducting a scoping review on ethics in physiotherapy is essential for consolidating current knowledge, identifying gaps, and guiding future research and practice. This review will provide a valuable resource for practitioners, educators, and policymakers seeking to enhance ethical standards and practices within physiotherapy.

### 1.2 Aim and Objective

This scoping review aims to answer the research question: “What is known about the intersection of ethics and physiotherapy?” The specific objectives are to (1) map the existing literature on ethical considerations in physiotherapy, including typical ethical dilemmas, adopted ethical principles, and the theoretical frameworks used to address these issues, (2) identify the methodologies employed in studying ethical principles and challenges in physiotherapy, and (3) highlight any gaps in knowledge regarding ethical considerations in this field.

## 2. Material and methsods

This scoping review will adhere to the methodological guidance for scoping reviews of the Joanna Briggs Institute (JBI) ^12^. The reporting will follow the Preferred Reporting Items for Systematic Reviews and Meta-Analyses extension for Scoping Reviews (PRIMA-ScR) ^13^.

### 2.1 Research team

The research team comprises four physiotherapists and one philosopher, all with qualitative and/or quantitative research backgrounds. One of the physiotherapists holds a master’s degree in philosophy, and another is pursuing a PhD in bioethics applied to rehabilitation. Additionally, one physiotherapist has a PhD in neuroscience and medical science, serving as a methodologist to oversee the methodological rigor of the process underlying this scoping review. The philosopher in the group has completed a PhD in bioethics. This diverse composition ensures that all relevant areas of expertise and knowledge necessary for this scoping review are comprehensively covered.

### 2.2 Eligibility criteria

Studies will be considered eligible for inclusion if they meet the Population, Concept and Context (PCC) framework criteria proposed by the JBI^12^.

#### 2.2.1 Population

We will include studies focusing on physiotherapists as professionals and physiotherapy as a discipline. This includes research on:

- **Physiotherapists**: Studies involving physiotherapists as practitioners, exploring their ethical challenges, decision-making processes, and professional conduct.
- **Physiotherapy Practice**: Research examining ethical issues related to the practice of physiotherapy, including interactions with patients, treatment methods, and the application of ethical principles in clinical settings.

In summary, the population of interest is limited to physiotherapists and the ethical aspects of their professional practice. Studies focusing on other healthcare professionals involved in physiotherapy will be excluded unless the ethical issues pertain specifically to physiotherapists. Similarly, research from the patient’s perspective will only be included if it directly examines physiotherapists’ ethical conduct or decision-making processes.

#### 2.2.2 Concept

The principal concept of interest is ethics and bioethics within the context of physiotherapy practice. Studies must address ethical issues, dilemmas, or principles as they pertain to physiotherapy and rehabilitation. Studies that focus solely on technical or procedural aspects of physiotherapy without addressing ethical dimensions will not be included. For instance, studies that examine only technical treatment methods or outcomes without considering their ethical implications will be excluded.

#### 2.2.3 Context

No specific restrictions will be applied to the context, as we intend to investigate studies from all geographical locations with participants regardless of specific demographic, social or cultural factors.

#### 2.2.4 Types of studies

All types of primary studies and publications (both qualitative and quantitative) will be included in this review with no restrictions to time, geographical location, setting and language. Reviews, editorials, conference abstracts, commentaries, expert opinions, letters to editors, book review chapters or study protocols will be excluded. However, their references will be checked for eligible studies.

### 2.3 Search strategy and information sources

The search strategy will involve the following databases: PubMed, Medline, Embase, Cochrane Central, Web Of science, CINAHL, PsychInfo, and Pedro. These databases were selected for their comprehensive coverage of health research and their ability to track citations across various disciplines. PubMed and Medline cover biomedical literature extensively, while Embase offers strong coverage of pharmacology and drug-related studies. Cochrane Central is crucial for systematic reviews and clinical trials, CINAHL covers nursing and allied health literature, PsychInfo includes psychological and behavioral studies, and Pedro focuses on evidence-based practice in physiotherapy.

A search string has been prepared for PubMed and will be adapted across all these databases. No limitations will be set on the search strategy or the study date (Supplementary File 1). The string will be converted to be used in all the other databases. These databases were selected due to their relevance to health research and their ability to track citations. No limitations will be set for the search strategy or the date of the study. A grey literature search will also follow the Canadian Agency for Drugs and Technologies in Health (CADTH) tool for searching health-related ^14^. The CADTH tool makes the grey literature searching process transparent and systematic ^15^. If required, authors will be contacted for further information or missing data. If needed, the search strategy will be modified and adapted to balance the relevance of the records following an interactive approach to scoping review. Any changes will be highlighted in the scoping review output. The International Prospective Register of Systematic Reviews database (PROSPERO) was consulted to check for ongoing reviews on this topic. No systematic reviews were found on this topic.

### 2.4 Study selection

All entries will be uploaded to Covidence (www.covidence.org), where duplicates will be automatically removed. The screening process will be conducted by two researchers (GB, FP) in the blind. A title and abstract review will be conducted, followed by a full-text screening. A pilot test, pre-formal screening for a random of 10% of records retrieved, will be conducted as a calibration exercise to improve reliability across reviewers. The formal screening will start if the percentage interrater agreement is >90%. Otherwise, the inclusion and exclusion criteria will be further specified, and another pilot test will be performed. In case of conflict, a third author will be consulted (SB). Reasons for the exclusion will be reported in the scoping review report. The final included studies will be mapped through the scoping review. A graphical representation of the selection of studies will be presented, adopting the Preferred Reporting Items for Systematic Reviews and Meta-analyses (PRISMA) flow diagram ^16^. The included studies will be uploaded to a OneDrive folder accessible to all team members. The studies’ authors will be contacted if we cannot find the full text of their papers.

### 2.5 Data extraction

Data will be charted based on the JBI Standardized Data Extraction Form ^17^. The following information will be extracted from the included studies:

- **Authors and year of publication**: Details about the authors and the year the study was published.
- **Country of origin**: The country or countries where the study was conducted.
- **Aims and purpose**: A description of the study’s aims and objectives.
- **Population and sample size**: Characteristics of the studied population and the sample size.
- **Study design**: The type of study conducted (e.g., qualitative, quantitative, review, etc.).
- **Ethical issues addressed**: Specific ethical dilemmas or issues discussed (e.g., patient autonomy, informed consent, professional conduct, confidentiality).
- **Domains of physiotherapy**: Areas of physiotherapy practice covered in the study (e.g., musculoskeletal rehabilitation, neurorehabilitation, rehabilitation techniques, patient interactions).
- **Outcomes**: Results related to the ethical issues (e.g., impact on patient care, professional conduct).
- **Methods of ethical assessment**: Tools or methods used to assess ethical issues (e.g., qualitative interviews, surveys, ethical frameworks).
- **Philosophical framework**: Theories or philosophical approaches used to analyze the ethical issues.
- **Conclusion**: A summary of how ethical issues were addressed and their implications for practice.

Any changes made to the data extraction form will be documented in the final scoping review. This form will be reviewed by all researchers involved and tested before implementation, following the same screening pilot test method. Two researchers (GB, FP) will independently extract the data. Given the iterative nature of the data extraction, other data may be added to the proposed draft. The modifications will be reported in the full scoping review.

### 2.6 Data synthesis

The results will be narratively synthesized to organize and classify the ethical issues and principles identified in the context of physiotherapy into overarching themes. This synthesis will involve grouping the findings into key thematic areas, such as ethical dilemmas in patient interactions, professional conduct, and ethical decision-making in various physiotherapy settings.

We will provide a descriptive summary of the findings, highlighting how ethical considerations are addressed across different aspects of physiotherapy practice. This summary will include identifying gaps in the literature where further research is needed and suggesting potential areas for future investigation.

All included studies will be reported and mapped to illustrate the breadth of the search and the data extracted. The results will be summarized in tables and graphs to visually represent the distribution of ethical topics and methodologies. Given the iterative nature of the scoping review process, additional categories or themes may be introduced as necessary to ensure a comprehensive analysis of the ethical dimensions in physiotherapy.

### 2.7 Methodological quality appraisal

No critical appraisal of the risk of bias will be performed in line with guidance on the scoping review ^12^, as we intend to map the available evidence rather than provide clinical and synthesised answers to a question.

## 3. Discussion

This scoping review aims to systematically explore and analyze the scientific literature on ethical issues within the field of physiotherapy. The primary objectives of this review are to (1) map the existing literature on ethical considerations in physiotherapy practice, (2) identify the methodologies used to assess these ethical issues, and (3) highlight any gaps in knowledge regarding the integration of ethical principles into physiotherapy.

We hypothesise that the review will reveal a concentration of studies focusing on specific ethical dilemmas such as patient autonomy, informed consent, and confidentiality, with potentially less attention given to broader ethical frameworks and their application across various aspects of physiotherapy practice. By outlining this protocol, we seek to provide a clear and systematic approach for conducting the review, minimizing potential reporting biases and improving the transparency of our work.

The protocol follows the methodological framework established for conducting scoping reviews ^12,13^. Any deviations from this protocol will be documented and addressed in the final scoping review report. The findings from this review will be disseminated through a peer-reviewed publication and presentations at relevant conferences to contribute to the understanding and development of ethical practices in physiotherapy.

## Supporting information

Supplemental file - researh string

## Data Availability

All data produced in the present work are contained in the manuscript

## Acknowledgements

None to declare.

## Ethics Statement

No Ethics Committee was needed for this study.

