## Supplemental file - researh string for "The Role of Ethics in Physiotherapy: A Scoping Review Protocol"

**Supplementary File 1**: Research String

| **Medline via Pubmed consulted in date 17/10/2024** | | | | |
| --- | --- | --- | --- | --- |
| **Population** | 1 | **Physical Therapy Modalities** | **[Mesh Term]** | 187,529 |
|  | 2 | **Physical Therapy Specialty** | **[Mesh Term]** | 3,087 |
|  | 3 | **Rehabilitation** | **[Mesh Term]** | 369,104 |
|  | 4 | Rehabilitation | [Title/Abstract] | 842,130 |
|  | 5 | Physiotherap* | [Title/Abstract] | 83,274 |
|  | 6 | "Physical therap*" | [Title/Abstract] | 123,518 |
|  | 7 | "Physical rehabilitat*" | [Title/Abstract] | 3,744 |
|  | 8 | "Rehabilitation therap*" | [Title/Abstract] | 4,778 |
|  | 9 | Physiotherapist | [Title/Abstract] | 5,056 |
|  | 10 | Physiotherapists | [Title/Abstract] | 8,527 |
|  | 11 | “Physical therapist” | [Title/Abstract] | 3,808 |
|  | 12 | “Physical therapists” | [Title/Abstract] | 6,165 |
|  | 13 | 1 OR 2 OR 3 OR 4 OR 5 OR 6 OR 7 OR 8 OR 9 OR 10 OR 11 OR 12 | | 571,211 |
| **Concept** | 14 | **Ethics** | **[Mesh Term]** | 158,840 |
|  | 15 | **Ethics Consultation** | **[Mesh Term]** | 1,435 |
|  | 16 | **Ethics, Research** | **[Mesh Term]** | 9,169 |
|  | 17 | **Codes of Ethics** | **[Mesh Term]** | 5,564 |
|  | 18 | **Principle-Based Ethics** | **[Mesh Term]** | 34,111 |
|  | 19 | **Ethics, Clinical** | **[Mesh Term]** | 65,200 |
|  | 20 | **Ethics, Professional** | **[Mesh Term]** | 75,373 |
|  | 21 | **Ethics, Nursing** | **[Mesh Term]** | 10,626 |
|  | 22 | **Ethics, Medical** | **[Mesh Term]** | 48,722 |
|  | 23 | **Ethical Theory** | **[Mesh Term]** | 3,678 |
|  | 24 | **Bioethics** | **[Mesh Term]** | 12,174 |
|  | 25 | Ethics | [Title/Abstract] | 99,889 |
|  | 26 | Bioethics | [Title/Abstract] | 17,089 |
|  | 27 | "Professional ethics" | [Title/Abstract] | 9,808 |
|  | 28 | “Ethical issue*” | [Title/Abstract] | 15,479 |
|  | 29 | "Ethical dilemma*” | [Title/Abstract] | 5,565 |
|  | 30 | "Ethical principle*" | [Title/Abstract] | 4,657 |
|  | 31 | "Moral obligation*" | [Title/Abstract] | 1,053 |
|  | 32 | "Moral issue*" | [Title/Abstract] | 789 |
|  | 33 | “Moral principle*” | [Title/Abstract] | 541 |
|  | 34 | "Moral dilemma*” | [Title/Abstract] | 1,333 |
|  | 35 | “Ethical decision-making” | [Title/Abstract] | 1,705 |
|  | 36 | “Moral decision-making” | [Title/Abstract] | 492 |
|  | 37 | "Ethical consideration*" | [Title/Abstract] | 8,319 |
|  | 38 | “Ethical challenge*” | [Title/Abstract] | 4,026 |
|  | 39 | “Moral consideration*” | [Title/Abstract] | 281 |
|  | 40 | “Moral challenge*” | [Title/Abstract] | 203 |
|  | 41 | "Bioethical issue*” | [Title/Abstract] | 510 |
|  | 42 | “Bioethical dilemma*” | [Title/Abstract] | 104 |
|  | 43 | 14 OR 15 OR 16 OR 17 OR 18 OR 19 OR 20 OR 21 OR 22 OR 23 OR 24 OR 25 OR 26 OR 27 OR 28 OR 29 OR 30 OR 31 OR 32 OR 33 OR 34 OR 35 OR 36 OR 37 OR 38 OR 39 OR 40 OR 41 OR 42 | | 239,332 |
|  | 44 | 13 AND 43 | | **5,907** |

**Context** was not applicable.
